# Electronic consent for school-age vaccinations: a rapid review

**DOI:** 10.1101/2025.05.15.25327678

**Authors:** Chukwudi Okolie, Golibe Ezenwugo, Mai Barry, Kerryann Richmond-Russell, Simon Cottrell

**Affiliations:** Public Health Wales Vaccine Preventable Disease Programme, Wales, United Kingdom; Public Health Wales Evidence Service, Wales, United Kingdom

## Abstract

**Background:** The use of digital or electronic processes for gathering consent to immunisation for school-age vaccinations (“e-consent”) has the potential to improve the efficiency of the process. However, little is known currently about the effectiveness of e-consenting or implications on vaccination uptake and equity.

**Methods:** MEDLINE, Embase, Cochrane Library, Scopus, PsycINFO, and Google Scholar were systematically searched for relevant publications from January 2000 until November 2024. Studies included in this review reported impact data on the use of digital or electronic processes as a means of obtaining informed consent for school-aged vaccinations. The quality of research included in the rapid review was assessed using the JBI critical appraisal tool for assessment of risk of bias for randomised controlled trials, the JBI checklist for quasi-experimental studies, and the mixed methods appraisal tool (MMAT). A narrative synthesis approach was used to analyse data and present findings.

**Results:** Four studies met the criteria for inclusion in the review. Included studies were conducted in the UK (n = 2) and USA (n = 2). Overall, the review findings appeared to suggest a preference by parents of school children for e-consent systems. Some parents found e-consent forms easy to use, however some others found the transition from traditional paper consent to e-consent difficult to adapt to. The use of e-consent did not appear to improve the timeliness of consenting or vaccination uptake. Language barriers, practical issues with transitioning to the new e-consent system, and concerns over lack of trust in electronic data, were among the reasons given for lack of engagement with the system.

**Conclusions:** There is currently growing interest on the use of e-consent as a means of obtaining informed consent for school-aged vaccinations. However, there is a paucity of high-quality evidence on the benefits and limitations of e-consent. None of the four studies included in this review identified that use of e-consent led to improved uptake, and none evaluated the impact of e-consent on vaccine equity. The impact of e-consent on overall level of consent/ non-consent return was variable. Transitioning from paper to e-consent systems has been difficult for some parents and adolescents to adapt to. Further evaluations of e-consent are required to inform its use in vaccination processes. Although utilisation of e-consent could yield efficiencies in gathering of consent, evidence that it leads to any benefit in vaccination outcomes is very limited.

## Introduction

Immunisation programmes around the world are increasingly incorporating vaccines that target age groups beyond infancy and early childhood, into their national immunisation schedules (World Health Organization, 2014). Educational settings play a vital role in the delivery of these programmes. Delivering immunisation programmes in schools makes vaccines more accessible to pupils, ensures timely protection and improved uptake (Crocker et al., 2012), and is an important opportunity to check that children are up to date with all their routine immunisations (UK Health Security Agency, 2024a). In the UK, vaccines routinely offered to children and young people in school include seasonal influenza vaccine, Human papillomavirus (HPV) vaccine, MenACWY vaccine, Td/IPV (3 in 1 booster), and the MMR check and offer (UK Health Security Agency, 2024b, Welsh Government, 2024).

Consent to immunisation is a legal requirement before vaccines can be administered to school children. For consent to immunisation to be valid, it must be voluntary and informed, and the person consenting must have the capacity to make the decision (Ramsay, 2021). This is usually done by the person themselves or someone with parental responsibilities. In high and middle-income countries, the most common method of collecting consent is by completion of a formal written consent form (Footer and Foster, 2022). However, logistical challenges involved in getting paper forms to and from parents, and a desire to save time, resources, and increase vaccination uptake, has led to calls for a more streamlined consent process (Chantler et al., 2019, Footer and Foster, 2022).

The use of digital tools and web-based technology for obtaining informed consent is not new and has become increasingly popular over the last decade. Digital or electronic consent (e-consent) systems have the potential to improve the efficiency of the consenting process and various vaccination-related outcomes. Recent studies on e-consent have shown promising results in various healthcare settings including, clinical care (Chimonas et al., 2023), medical research (Gesualdo et al., 2021), and neurosurgery (Bethune et al., 2018). E-consent systems for school-age vaccinations are currently being trialled in several NHS trusts in the UK. At present, little is known about the effectiveness of these systems, whether they have any impact on vaccination-related outcomes, or any potential risks to digital equity.

This rapid review therefore seeks to assess the evidence on the use of digital or electronic consent (e-consent) as a means of obtaining informed consent for school-aged vaccinations, in comparison to traditional paper-based consent processes.

## Objectives

The objectives of this rapid review are to:

1. Identify, review, and synthesise the findings from published articles describing e-consent systems for obtaining informed consent for school-aged vaccinations.
2. Evaluate the effectiveness of e-consent over paper-consent processes in terms of timeliness and completeness of consent return, uptake and equity of vaccination.

## Methods

This rapid review was based on abbreviated systematic review methods and conducted in compliance with PRISMA guidelines (Page et al., 2021).

### Search strategy

Six electronic databases – MEDLINE, Embase, Cochrane Library, Scopus, PsycINFO, and Google Scholar – were systematically searched on 27 November 2024 for relevant publications. A comprehensive search strategy (Appendix 1), developed in MEDLINE and adapted for use in each of the other databases, was used for the searches. Database subject and keyword searches were conducted using a range of terms representing “school”, “child”, “electronic consent”, and “vaccination”. Searches were limited to literature from OECD countries (pre-1974 membership) published in English language from 2000 onwards. A 2000 date limit was agreed as this would be more likely to capture literature on digital or electronic consenting processes. The reference lists of included studies were also reviewed for relevant publications. All search results were exported into an EndNote library (version 20) and duplicates removed. The review protocol is available in the International Prospective Register of Systematic Reviews (registration number: CRD42025637787).

### Criteria for considering studies for the review

Studies eligible for inclusion in this review reported effectiveness data on the use of digital or electronic consent as a means of obtaining informed consent for school-aged vaccinations. This included both secondary and primary research studies. The complete eligibility criteria for inclusion of studies in this review can be found in Table 1.

**Table 1:** Eligibility criteria.

|  | Inclusion criteria | Exclusion criteria |
| --- | --- | --- |
| Population | School-age children, i.e. children and young people in school years reception to year 11 (ages 4 to 16), parents/carers of school-age children, parent-child dyads | Non-school age children, adults |
| Intervention | Digital/e-consent forms for school-age vaccinations (seasonal flu, HPV, DPT, MenACWY, MMR)<br>• Accessed via smart phone, tablet, computer | Non-digital consent forms |
| Comparator | Paper-based consent |  |
| Outcomes | Efficiency/effectiveness of return of consent/non-consent, vaccination uptake, acceptability (parental, child, staff), equity |  |
| Study design | Secondary research, comparative quantitative primary studies, qualitative studies |  |
| Countries | OECD countries pre-1974 | Non-OECD countries or post-1974 membership |
| Language of publication | English language | Non-English language |
| Publication date | 2000 - 2024 | Pre - 2000 |
| Publication type | Published and preprint (preprint only if limited published research identified) | Commentaries, editorials, letters, conference abstracts |

### Selection of studies for inclusion in the review

Following removal of duplicates in Endnote, all articles were imported into the systematic reviewing software Rayyan (Ouzzani et al., 2016) for screening. A two-stage screening process was undertaken by four independent reviewers (CO, GE, MB, & KRR). First, all reviewers screened titles and abstracts against the eligibility criteria in Table 1, to identify potentially eligible publications. In the second stage, all reviewers independently assessed full texts of eligible studies to identify studies to be included in the review. The reviewers also examined the reference lists of all included studies to identify other potentially eligible studies. Any disagreements at either stage were resolved by consensus.

### Critical appraisal

The quality of included studies was assessed using three critical appraisal tools – the JBI critical appraisal tool for assessment of risk of bias for randomised controlled trials (Barker et al., 2023), the JBI checklist for quasi-experimental studies (Barker et al., 2024), and the mixed methods appraisal tool (MMAT) (Hong et al., 2018). Two reviewers (CO & GE) independently assessed the methodological quality of all included studies. Any disagreements were resolved by consensus.

### Data extraction

Two reviewers (CO & GE) independently extracted data from included studies using a data extraction table. For all included studies, data were extracted on general information (study reference, including title, authors, year of publication, country), study details (study design, aims, populations, sample size, setting, intervention details, outcome measures), and study findings. Study authors’ conclusions and any additional comments of note were also extracted. Both reviewers compared all extracted data and resolved any disagreements by discussion.

### Data analysis and synthesis

A narrative synthesis approach was used for evidence synthesis. The use of meta-analysis to synthesise quantitative findings was considered, however, due to the small number of eligible studies, the limited data provided, and the different outcomes measured, it was not practical to conduct a valid meta-analysis.

## Results

A total of 3756 citations were identified through electronic database searching. After deduplication, the initial number of citations decreased to 2878. Of these, 2865 citations were excluded after screening of titles and abstracts. Thirteen articles were assessed at full-text for eligibility, of which four articles met the criteria for inclusion in the review. The study selection process is shown in Figure 1.

**Figure 1:**
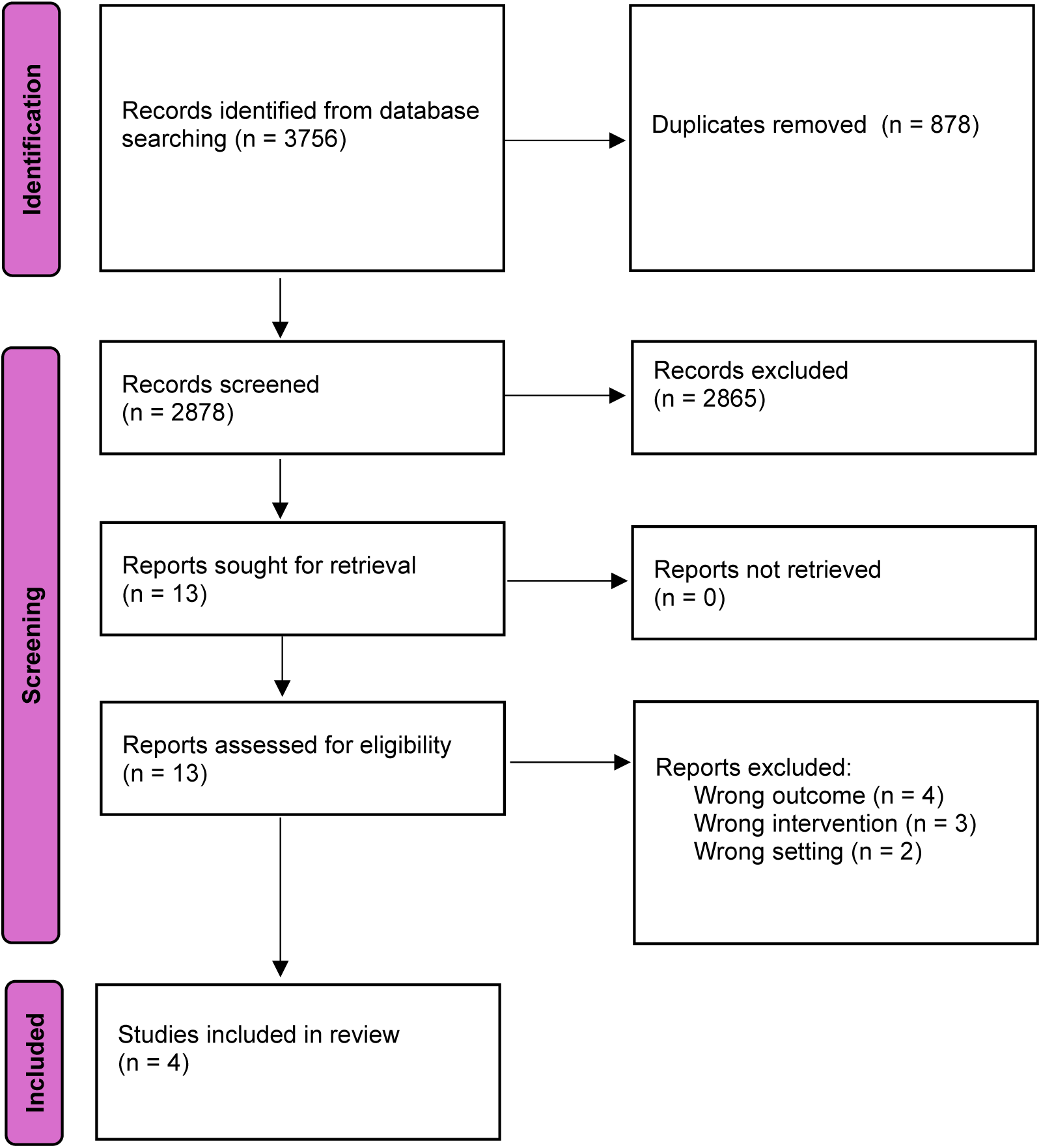
PRISMA flow diagram.

### Characteristics of included studies

Of the four included studies, two were cluster randomised controlled trials (RCTs), one was a quasi-experimental study, while the remaining study was a mixed methods study. The included studies were conducted in the UK (n = 2) and USA (n = 2). Three of the four included studies were conducted in secondary schools and targeted adolescent school children, while one study was conducted in elementary schools. Two studies (Szilagyi et al., 2016, Szilagyi et al., 2018) focused on school-located influenza vaccinations, while the other two studies (Chantler et al., 2020, Footer and Foster, 2022) focused on HPV vaccination. Outcomes reported in included studies included consent form return rates (Chantler et al., 2020, Footer and Foster, 2022), vaccination uptake (Chantler et al., 2020, Footer and Foster, 2022, Szilagyi et al., 2016), outcome of consent (Chantler et al., 2020), and user satisfaction, preference, and acceptability (Chantler et al., 2020, Footer and Foster, 2022, Szilagyi et al., 2016, Szilagyi et al., 2018). Two related RCTs (Szilagyi et al., 2016, Szilagyi et al., 2018) investigated the impact of school-located influenza vaccinations, with an exploratory aim of assessing the use of web-based consent. The details of the individual studies can be found in Table 2.

**Table 2:** Characteristics of included studies.

| Citation (Country) | Study Details | Participants & setting | Key findings | Conclusions |
| --- | --- | --- | --- | --- |
| Chantler et al (2020). Does electronic consent improve the logistics and uptake of HPV vaccination in adolescent girls? A mixed-methods theory informed evaluation of a pilot intervention. BMJ open, 10(11), e038963.<br>**Country:** United Kingdom | **Study design:** Mixed methods (Qualitative and quantitative)<br>**Study aim:** To evaluate the usability and acceptability of an electronic consent (e-consent) pilot intervention for school-based immunisations and assess its impact on consent form returns and human papilloma virus (HPV) vaccine uptake.<br>**Intervention:** The e-consent intervention consisted of an online portal with an e-consent form and a data platform and related implementation procedures. An electronic consent portal where parents could record whether they agreed to or declined vaccination, and nurses could access data to help them manage the immunisation programme.<br>**Comparator:** Paper consent forms<br>**Data collection method and dates:** Data collection occurred in June – December 2018 (year 1 i.e., the pilot study which involved observations of HPV immunisation sessions, interviews with parents, adolescents, and staff, and feedback forms from schools) and June – July 2019 (year 2 which were qualitative follow-ups to evaluate adaptations after Year 1). The first period coincided with and followed the first year of e-consent | **Sample size:** Twenty-eight schools (14 paper and 14 e-consent schools) comprising of 3219 girls (1733 in paper consent and 1486 in e-consent schools) were included in the study. Twenty six of the 28 schools were state and two were private schools.<br>**Participants:** Participants included nurses and healthcare staff who administered the vaccination sessions and handled the consent process, data managers, school-link staff who organised the vaccination session and consent logistics, parents who were responsible for providing consent either electronically or using paper forms, and adolescent girls eligible for the first dose of the HPV vaccine.<br>**Setting:** The intervention was piloted in 14 secondary schools in seven London boroughs in 2018, with 14 other schools used as controls. | **Primary findings**<br>**Quantitative**<br>**Return of consent form:** 83% of consent forms (paper or e-consent) were returned prior to the vaccination session. However, among the 22 matched schools where this data was available, compared with paper schools timely (prior to the planned session) return was lower in the e-consent schools (73.3% vs 91.6%, $p=0.008$).<br>**Outcome of consent:** There was no statistically significant difference in the proportion of pupils for whom a 'yes' consent was received, prior to or on the day of the session, between the paper ($n=14$) and e-consent ($n=14$) schools (85% in e-consent schools, 83% in paper consent schools, $p=0.89$).<br>**Vaccination uptake:** There was no statistically significant difference in the proportion of pupils that were vaccinated at the scheduled vaccination session between the paper ($n=14$) and e-consent ($n=14$) schools (80.6% vs 81.3%, $p=0.93$).<br>The final vaccine uptake across all the schools was over 86%.<br>**Additional findings:**<br>**Qualitative**<br>**Resources and training:** Immunisation team members were positive about the development of the e-consent intervention but expressed reservations about not being able to review paper consent forms prior to immunisation. Due to tight deadlines only one orientation session took place before the e-consent intervention was introduced, which meant that the bulk of learning happened on the job. | **Authors' conclusions:** The pilot e-consent intervention did not improve consent form return or vaccine uptake due to challenges encountered in transitioning to new working practice. New technologies require embedding before they become incorporated in everyday practice. A re-evaluation once stakeholders are accustomed with electronic consent may be required to understand its impact.<br>**Study strengths and limitations:**<br>According to the study authors, key limitations include the lack of interviews with school staff to complement the feedback forms and not being able to access and interview parents who found the intervention more challenging to use.<br>This evaluation was limited to one cycle of implementation, which did not allow the authors to account for the time it |
|  | implementation, the second occurred during the use of e-consent in a different subset of schools.<br>**Outcome measures:**<br>• Return rates of consent form<br>• Outcome of consent<br>• HPV vaccination uptake<br>• Usability and acceptability of the e-consent system among parents, adolescents, and staff. |  | **Using the e-consent intervention:** The parents interviewed found the system easy to use and usually completed the form as soon as they received it. A few parents would have liked an email confirmation after they had completed the e-consent form. According to feedback from nurses and schools, not all parents found the intervention easy to access or use. Language barriers accounted for some difficulties, but practical issues also played a role.<br>**Transition: adapting to change and iterative development:** The initial 'buzz' about the e-consent intervention decreased over time. While some staff remained positive and receptive to the implementation coordinators enthusiasm and vision, others expressed a sense of half-heartedness about having to adapt quickly from a known way of obtaining consent, although with flaws (e.g., cost of paper, mileage clocked up in collecting paper consent forms from schools), to a new technology enabled way with some functional limitations in year 1. | takes for new practices to become embedded. The number of schools who declined to pilot the intervention was also not recorded.<br>**Notes/comments:**<br>This study used a 'theory of change' as an evaluation framework.<br>As the pilot and implementation phase were done within a year, the e-consent system was not fully operational. As a result, there was some reliance on e-consent responses being completed manually and this could have introduced some level of bias. |
| Footer & Foster (2022). The introduction of electronic consent for the school aged immunization program. Public Health Nursing, 39(1), 320-325.<br>**Country:**<br>United Kingdom | **Study design:** Quasi-experimental study<br>**Study aim:** To design and implement a more efficient and cost-effective process for gaining consent for school-aged immunisation, aligning with the goals of a "paperless National Health Service" and improve vaccine uptake.<br>**Intervention:** An electronic consent (eConsent) form for the routine school aged immunisation programme. The eConsent solution consisted of a set of SQL databases, used to collate consent web form submissions made by parents of schools age children for multiple different vaccinations. | **Sample size:** During the pilot phase in 2019 – 2020, 25 schools were involved with 2,983 e-consent forms rolled out. In the 2020-2021 countywide rollout, 9,496 pupils were eligible for vaccination with 6,937 e-consent forms returned (73.0% return rate).<br>**Participants:**<br>School-aged children aged 12 – 13 years old who were eligible for routine HPV1 immunisations, parents or guardians who provided consent using either electronic or traditional paper method and healthcare staff involved in organising and administering the vaccinations, handling the logistics and managing consent data. | **Primary findings**<br>**Impact on Vaccine Uptake:** During the pilot phase in 2019, vaccine uptake remained stable despite the introduction of a new system, with similar consent rates as previous years using paper forms. HPV1 uptake 2018-2019 (paper consent) = 89.6% HPV1 uptake 2019-2020 (eConsent) = 89.9%<br>However, vaccine uptake dropped by 20% in 2020–2021, which was attributed to external factors like COVID-19 disruptions and misinformation about vaccines.<br>HPV1 uptake 2020-2021 (eConsent) = 70.2%<br>**Consent form return rate:** With an average return rate of 80%, the e-consent form return rate was significantly higher than paper consent form returns. | **Author's conclusions:** The authors concluded that despite some setbacks, the e-consent system offers a significant opportunity to improve participant experience, reduce resource demands, and enhance healthcare delivery. They advocate for further use and data collection to establish its long-term impact.<br>**Study strengths and limitations**<br>**Stakeholder collaboration:** |
|  | **Comparator:** Traditional paper consent forms previously used for gaining parental consent for immunisation.<br>**Data collection method and dates:** Data collection involved tracking consent returns and vaccination uptake through the e-consent system during its pilot phase in the summer of 2019 and the subsequent launch in September 2020. Feedback was also gathered from stakeholders, including schools, parents, and immunisation teams, as well as performance data from the system.<br>**Outcome measures:**<br>Primary: Immunisation uptake rates and consent return rates before and after e-consent implementation.<br>Secondary: Efficiency improvements, resource savings (time and cost), and stakeholder satisfaction with the new system. | **Setting:**<br>The pilot intervention took place in 25 schools during the summer of 2019, and expanded countywide to all schools in Northamptonshire in September 2020. | There was also a significantly quicker return rate for e-consent forms compared to paper returns.<br>**User satisfaction:** Parents found it easier to engage with the service electronically, resulting in an improvement in the number of returned consent forms within the required period of time during the pilot.<br>**Stakeholder resistance:** Some schools, immunisation team staff, and parents showed resistance towards moving away from the traditional practice of paper consent forms, whereas others were keen to embrace change. Staff reported feelings of uncertainty and expressed a lack of trust in electronic data.<br>**Language and accessibility Issues:** Some parents, particularly those whom English is not their first language, struggled with the new process, thus prompting the addition of a language picker function. | There was close collaboration with stakeholders during development ensuring that the e-consent process was user-friendly and met service needs.<br>**Implementation challenges**<br>Technical issues with email communication and parents missing submission deadlines were highlighted as limitations.<br>**Notes/comments**<br>This study used a 'theory of change' as an evaluation framework.<br>There is limited data analysis on other potential factors that could have affected the reduction in consent return rates after phase 1. Likewise, there is limited qualitative feedback provided from the parents and schools on the resistance the authors/implementers encountered in phase 2 with the e-consent system. |
| Szilagyi et al (2016). School-located influenza vaccinations: a randomized | **Study design:** Cluster RCT<br>**Study aim:** To assess:<br>1. The impact of school-located influenza vaccination (SLIV) on overall influenza vaccination rates for elementary school children. | **Sample size:** 19776 children from 44 schools<br>**Participants:** Elementary school children (kindergarten to sixth grade) | **Primary findings:**<br>**Impact of SLIV on Influenza Vaccination:** Children in SLIV schools had higher influenza vaccination rates than children in control schools county-wide (54.1% vs 47.4%, $P < 0.001$), in suburban schools (61.9% vs 53.6%, $P < 0.001$), and in urban schools (43.9% vs 39.2%; $P < 0.001$). | **Authors' conclusions:** SLIV raised seasonal influenza vaccination rates county-wide and in both suburban and urban settings. SLIV did not substitute for primary care vaccinations in |
| trial. Pediatrics, 138(5).<br>**Country:**<br>USA | 2. The impact of SLIV in suburban and urban school districts<br>3. The impact of SLIV on vaccination in primary care offices<br>4. The use of Web-based informed consent by parents.<br>**Intervention:** Parents of children at SLIV schools were sent information and vaccination consent forms via e-mail, backpack fliers, or both (depending on school preferences) regarding school vaccine clinics. This included a web-based application for parental consent for influenza vaccination, and a tracking system to remind consented parents about upcoming vaccine clinics<br>**Comparator:** Parents of children at control schools received usual care<br>**Data collection method and dates:** Child names and birthdates were matched with the state immunisation registry - New York State Immunization Information System (NYSIIS) records to obtain influenza vaccination data for the current (2013-2014) and previous seasons.<br>Parental use of online consents (aim 4) was assessed by examining the online forms.<br>**Outcome measures:**<br>1. The primary outcome measure was receipt of $\geq 1$ influenza vaccination (from any location) during the study year ("vaccination anywhere").<br>2. Parental use of web-based consent | **Setting:** Elementary schools in six suburban and one urban school districts in Monroe County, New York | On bivariate analysis across the full study sample, students in SLIV schools had higher odds of receiving an influenza vaccination than students in control schools (OR = 1.28, [CI]: 1.20–1.38). SLIV was associated with increased vaccination rates in both suburban (OR = 1.34, CI: 1.23–1.47) and urban schools (OR = 1.21, CI: 1.08–1.35). From the multivariate analysis for suburban and urban districts combined, the odds of vaccination were higher among students attending SLIV schools than among students attending control schools (OR = 1.32, CI: 1.23–1.42). On stratified analysis, vaccination rates were higher among SLIV schools in both suburban districts (OR = 1.38, CI: 1.27–1.51) and the urban district (OR = 1.23, CI: 1.10–1.38).<br>**Use of Web-based Consent**<br>**Suburban schools:** Among the 5616 children in suburban SLIV schools, 3478 families received paper notifications only (62%); 239 (7%) consented for SLIV; and 113/239 (47%) consented online indicating that parents accessed the Web site noted on the paper notifications. Among the 1702 families who received e-mail notifications only, 66 (4%) consented for SLIV; 59/66 (89%) consented online; and 7 (11%) printed out and returned written consent forms. Among the 436 children notified both ways, 31 (7%) consented for SLIV; and 22/31 (71%) consented online.<br>**Urban schools:** Among the 4344 children at urban schools allocated to SLIV, all children received paper notifications only, 443 (10.2%) consented for SLIV, and 53/443 (12%) consented online.<br>**Impact of consent type on SLIV**<br>Type of consent (paper or Web-based) did not appear to markedly affect the impact of SLIV<br>**Likelihood of influenza vaccination as a function of notification type** | suburban settings where pediatricians often preorder influenza vaccine but did substitute somewhat in urban settings.<br>**Study strengths and limitations:** According to the study authors, study strengths included<br>1. A robust randomised controlled trial study design to measure impact of SLIV<br>2. Multiple school districts that enhanced generalizability<br>3. Large numbers of children included<br>4. Accurate assessment of influenza vaccination via the state immunisation registry<br>Study limitations included:<br>1. The study was conducted in a single county that may not be representative of other counties.<br>2. SLIV and control schools had slightly different baseline characteristics<br>3. NYSIIS may have lacked influenza vaccinations for children who immigrated to NYS just before the study<br>4. The study was resource-intensive. |
|  |  |  | **Suburban schools:** Vaccination rates were paper notification only (58% vaccinated), e-mail notification only (70%), or both paper and e-mail notification (63%), compared with control (54%). Utilising multilevel logistic regression with matched pair fixed effects and school random effects, and controlling for the previous year's influenza vaccination to adjust for the propensity to be vaccinated, the odds of receiving a vaccination in the current year were higher for paper only notification (OR = 1.42, $P < 0.001$) and e-mail-only notification (OR = 1.38, $P < 0.001$) but not for paper and e-mail combined notification (OR = 1.12, $P = 0.43$).<br>**Urban schools:** Paper only notification was associated with a 44% vaccination rate compared with 39% for controls (OR = 1.23, $P < 0.001$). | **Notes/comments:** The use of web-based consent was sought as an exploratory aim in this study. |
| Szilagyi et al (2018). School-located influenza vaccinations for adolescents: a randomized controlled trial. Journal of Adolescent Health, 62(2), 157-163.<br>**Country:**<br>USA | **Study design:** Cluster RCT<br>**Study aim:**<br>1. To evaluate the effect of school-located influenza vaccination (SLIV) on adolescents' influenza vaccination rates.<br>2. To estimate the degree to which SLIV for adolescents substituted for vaccination in primary care<br>3. To assess the use of a novel web-based vaccine consent process to offer parents the option of consenting online for adolescent SLIV or printing forms for written consent<br>**Intervention:** School located influenza vaccination (SLIV)<br>**Comparator:** Usual care<br>**Data collection method and dates:** After the vaccination season (April 1, 2016), adolescent names and birthdates were matched with NYSIS records to obtain | **Sample size:** 17,650 students from 20 schools<br>**Participants:** Adolescents attending middle and high school<br>**Setting:** Eight suburban and two urban schools in Monroe County, New York | **Primary findings:**<br>**Impact of school-located influenza vaccination on overall influenza vaccination rates:** The overall influenza vaccination rate among students attending SLIV schools ("vaccinated anywhere") was higher than rates among students attending control schools in both suburban schools (51.2% vs. 45.8%, $p < 0.001$) and urban schools (36.7% vs. 31.4%, $p = 0.01$). In multivariate analysis, among suburban schools, the odds of an adolescent receiving an influenza vaccination anywhere was higher for adolescents attending SLIV schools than adolescents attending control schools (odds ratio [OR] 1.27, 95% confidence interval [CI] 1.18–1.38). However, among urban schools, adolescents at SLIV schools did not have statistically higher vaccination rates than adolescents at control schools.<br>**Use of web-based consent (exploratory aim)**<br>Of the 65 students enrolled in SLIV at the three schools exclusively using e-mail notifications, 37 (57%) consented online, whereas 28 (43%) parents printed out, signed, and returned completed paper consent forms to school. Among the 188 adolescents attending the five schools that sent both e-mail and paper | **Authors' conclusions:**<br>1. SLIV for adolescents improved influenza vaccination rates by a modest amount within suburban schools but not within urban schools.<br>2. SLIV for adolescents did not substitute for vaccines delivered in primary care.<br>3. Web-based consent for SLIV is now a viable option for SLIV programmes targeting adolescents.<br>**Study strengths and limitations:** According to the study authors, strengths of this study include:<br>1. The use of an RCT design to evaluate the impact of SLIV |
|  | influenza vaccination data for the past three influenza vaccination seasons.<br>**Outcome measures:** The primary outcome was receipt of an influenza vaccination anywhere during the 2015–2016 study year.<br>To assess the exploratory aim of parent selection of web-based consent (vs. printing off consent forms) at suburban schools, an assessment of the rates of web-based consent and influenza vaccination with different notification methods utilizing multilevel logistic regression, was done. |  | notifications, 152 (81%) consented online ($p < 0.001$ compared with e-mail-only notifications) and 36 (19%) returned completed paper consent forms. | 2. The inclusion of multiple school districts to raise generalizability.<br>3. Large numbers of adolescents, at least in suburban schools.<br>4. An assessment of receipt of influenza vaccination via NYSIIS which is mandated in NYS, and thus were able to assess location of vaccination and evaluate substitution of SLIV for vaccinations received elsewhere.<br>Limitations include:<br>1. The trial was conducted in one diverse county which may not be representative of other counties.<br>2. Power to detect differences for urban schools was low.<br>3. NYSIIS may have lacked records of influenza vaccinations for children who moved to NYS just before the study<br>**Notes/comments:** Szilagyi 2016 and Szilagyi 2018 are related trials, however they differ in their target population groups (they assess outcomes in elementary school children and middle/high school students respectively). |
|  |  |  |  | The use of web-based consent was sought as an exploratory aim in this study, and as a result the study may have been underpowered to detect any change. The study did not directly measure the impact of electronic vs paper consent |

### Quality assessment

Due to the variation in study designs of the individual studies, quality was assessed using three critical appraisal tools – the JBI critical appraisal tool for assessment of risk of bias for randomised controlled trials (Barker et al., 2023), the JBI checklist for quasi-experimental studies (Barker et al., 2024), and the mixed methods appraisal tool (MMAT) (Hong et al., 2018). The two RCTs were judged to be of moderate quality, the mixed methods study was judged to be of good quality, while the quasi-experimental study was judged to be of very low quality, due to limited information on the study sample and statistical techniques used in the study. Critical appraisal details can be seen in Tables 3, 4, and 5.

**Table 3:**
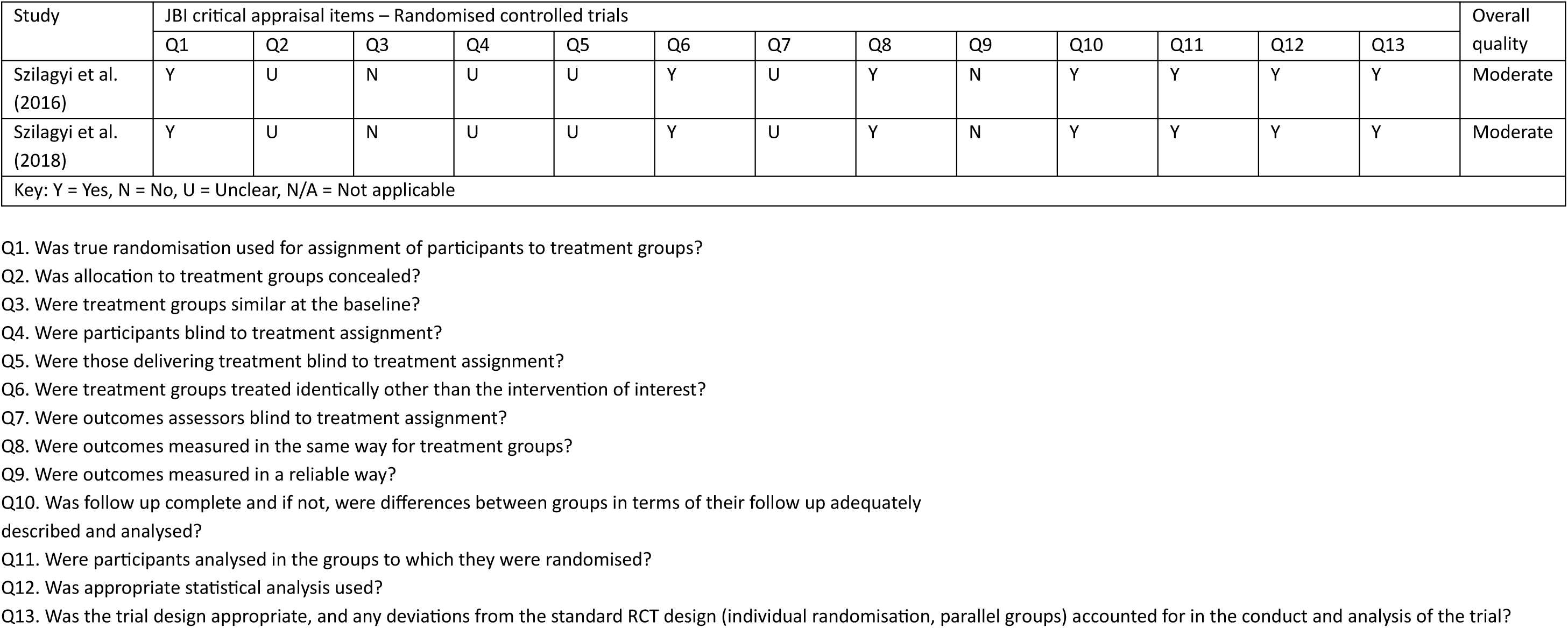
Critical appraisal (RCTs)

| Study | JBI critical appraisal items – Randomised controlled trials |  |  |  |  |  |  |  |  |  |  |  |  | Overall quality |
| --- | --- | --- | --- | --- | --- | --- | --- | --- | --- | --- | --- | --- | --- | --- |
|  | Q1 | Q2 | Q3 | Q4 | Q5 | Q6 | Q7 | Q8 | Q9 | Q10 | Q11 | Q12 | Q13 |  |
| Szilagyi et al. (2016) | Y | U | N | U | U | Y | U | Y | N | Y | Y | Y | Y | Moderate |
| Szilagyi et al. (2018) | Y | U | N | U | U | Y | U | Y | N | Y | Y | Y | Y | Moderate |
| Key: Y = Yes, N = No, U = Unclear, N/A = Not applicable |  |  |  |  |  |  |  |  |  |  |  |  |  |  |

Q1. Was true randomisation used for assignment of participants to treatment groups?
Q2. Was allocation to treatment groups concealed?
Q3. Were treatment groups similar at the baseline?
Q4. Were participants blind to treatment assignment?
Q5. Were those delivering treatment blind to treatment assignment?
Q6. Were treatment groups treated identically other than the intervention of interest?
Q7. Were outcomes assessors blind to treatment assignment?
Q8. Were outcomes measured in the same way for treatment groups?
Q9. Were outcomes measured in a reliable way?

**Table 4:** Critical appraisal (Quasi-experimental studies)

| Study | JBI critical appraisal items – Quasi-experimental studies |  |  |  |  |  |  |  |  | Overall quality |
| --- | --- | --- | --- | --- | --- | --- | --- | --- | --- | --- |
|  | Q1 | Q2 | Q3 | Q4 | Q5 | Q6 | Q7 | Q8 | Q9 |  |
| Footer & Foster (2021) | Y | Y | U | Y | N | Y | U | N | U | Very low |
| Key: Y = Yes, N = No, U = Unclear, N/A = Not applicable |  |  |  |  |  |  |  |  |  |  |

Q1. Is it clear what is the cause and what is the effect?
Q2. Was there a control group?
Q3. Were the participants included in any comparisons similar?
Q4. Were the participants included in any comparisons receiving similar treatment/care, other than the exposure or intervention of interest?
Q5. Were there multiple measurements of the outcome both pre and post the intervention/exposure?
Q6. Were the outcomes of participants included in any comparisons measured in the same way?
Q7. Were outcomes measured in a reliable way?
Q8. Was follow up complete and if not, were differences between groups in terms of their follow up adequately described and analysed?
Q9. Was appropriate statistical analysis used?

**Table 5:**
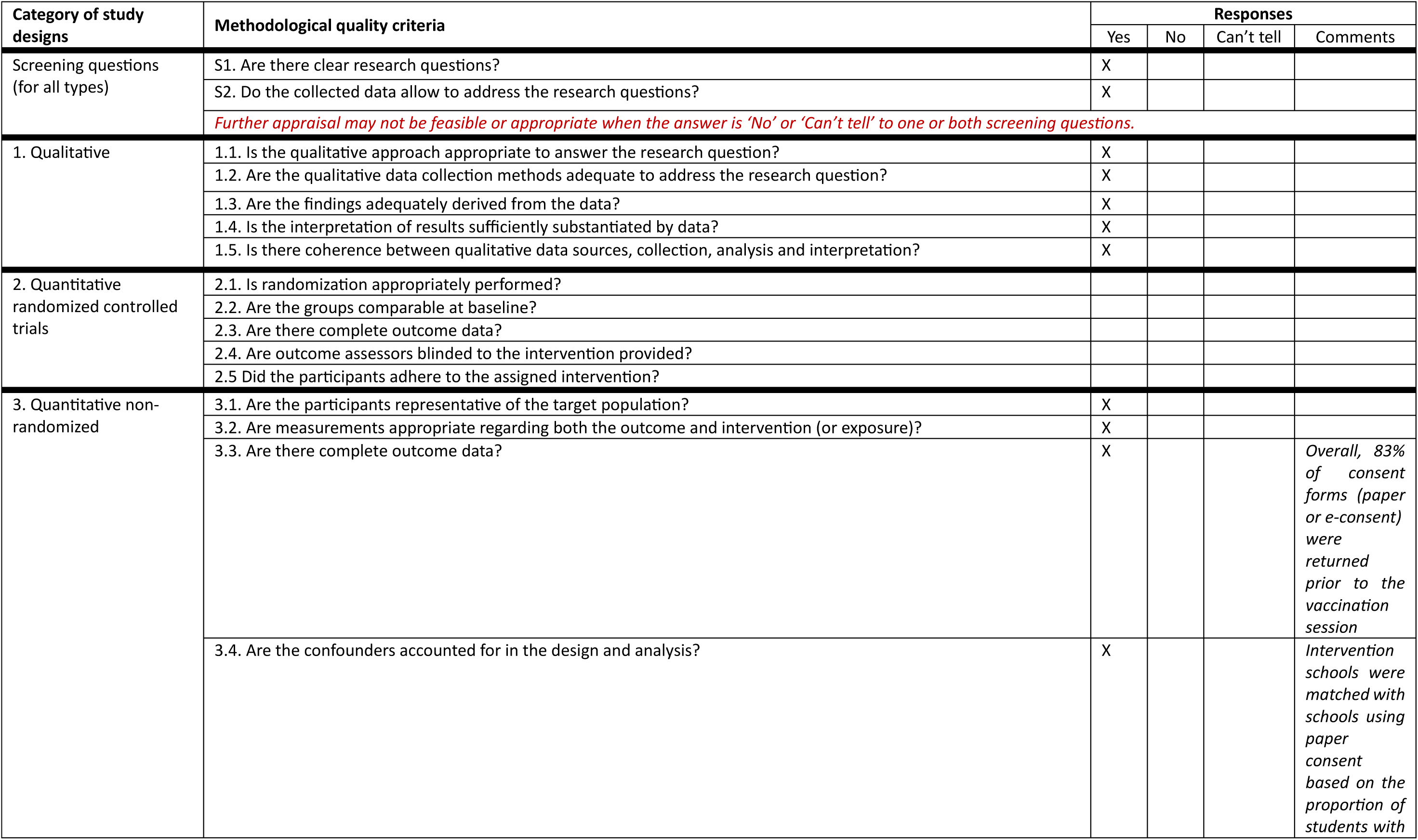
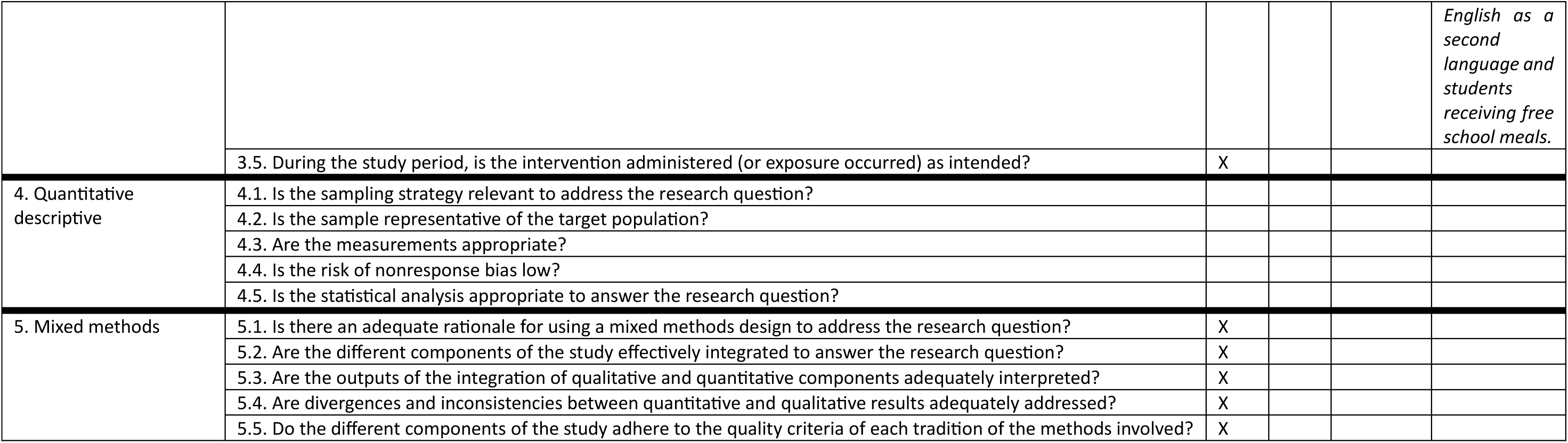
Critical appraisal (Mixed-methods)

## Study findings

### Impact of e-consent on consent form return

Two studies assessed the impact of e-consent on consent form return (Chantler et al., 2020, Footer and Foster, 2022). Study findings were mixed.

Chantler et al., (2020) conducted a mixed-methods study to evaluate the usability and acceptability of an electronic consent pilot intervention for school-based immunisations, and assessed its impact on consent form returns and HPV vaccine uptake. The study reported that timely return was lower in the e-consent schools (73.3% vs 91.6%, p=0.008) compared with schools issued paper consent forms. Reasons for lower consent form return in e-consent schools included difficulties encountered by some parents in accessing and using the intervention.

Footer and Foster (2022) piloted an e-consent form with a cohort of 12–13-year-olds eligible for the HPV1 vaccine, and compared outcomes with children who had been issued with the traditional paper consent form. The study reported an average e-consent return rate of 80% which was significantly higher than the paper consent return rate. There was also a significantly quicker return rate for e-consent forms compared to paper returns.

### Impact of e-consent on vaccination uptake

Three studies assessed the impact of e-consent on vaccination uptake (Chantler et al., 2020, Footer and Foster, 2022, Szilagyi et al., 2016).

Chantler et al., (2020) assessed the impact of an e-consent intervention on HPV vaccination uptake and found that there was no statistically significant difference in the proportion of pupils that were vaccinated at the scheduled vaccination session between the paper (n=14) and e-consent (n=14) schools (80.6% vs 81.3%, p=0.93).

Footer and Foster (2022) reported that during the pilot phase of an e-consent form for the routine school aged immunisation programme, vaccine uptake remained stable - with similar consent rates as the previous year’s using paper forms; HPV1 uptake 2018-2019 (paper consent) = 89.6% vs. HPV1 uptake 2019-2020 (e-consent) = 89.9%. However, vaccine uptake dropped by 20% the following year, HPV1 uptake 2020-2021 (e-consent) = 70.2%. This decrease in consent was attributed to external factors like COVID-19 disruptions and misinformation about vaccines.

Szilagyi et al., (2016) assessed the impact of offering school-located influenza vaccination (SLIV) clinics using both web-based and paper consent upon overall influenza vaccination rates among elementary school children. The study findings reported that type of consent (paper or web-based) did not appear to markedly affect the impact of SLIV (no data provided).

### Impact of e-consent on outcome of consent

One study assessed the impact of the introduction of an e-consent intervention on outcome of consent (Chantler et al., 2020). This study compared e-consent schools with paper consent schools in terms of outcome of consent, and reported no statistically significant difference in the proportion of pupils for whom a ‘yes’ consent was received between the paper (n=14) and e-consent (n=14) schools (85% in e-consent schools, 83% in paper consent schools, p = 0.89).

### User satisfaction, preference, and acceptability

Four studies reported user satisfaction, preference, or acceptability outcomes (Chantler et al., 2020, Footer and Foster, 2022, Szilagyi et al., 2016, Szilagyi et al., 2018).

Chantler et al., (2020) used qualitative methods to capture data on the usability and acceptability of e-consent and paper consent from school staff, parents, and adolescents. These methods included the use of school feedback forms and interviews.

The parents interviewed found the e-consent system easy to use and usually completed the form as soon as they received it. According to feedback from nurses and schools, not all parents found the intervention easy to access or use. Language barriers and practical issues with the system, accounted for some of these difficulties. Some staff also found the transition from paper to e-consent difficult to adapt to, which they felt removed some level of control from schools.

Footer and Foster (2022) reported that parents found it easier to engage with the programme electronically, which resulted in an improvement in the number of returned consent forms. However, it was found that some schools, immunisation team staff, and parents were resistant towards moving away from traditional paper consent forms, citing concerns over lack of trust in electronic data. Some parents also struggled with this new process particularly those whom English is not their first language.

Szilagyi et al., (2016) reported that among parents who consented for school-located influenza vaccination (SLIV) clinics, nearly half (47%) of those notified by backpack fliers and four-fifths (89%) of those notified by e-mail consented online, indicating a preference for web-based consent.

Szilagyi et al., (2018) assessed the use of a novel web-based vaccine consent process to offer parents the option of consenting online for adolescent school-located influenza vaccination (SLIV) clinics or printing forms for written consent. Study findings showed that of the 65 students enrolled in SLIV at the three schools exclusively using e-mail notifications, 37 (57%) consented online, whereas 28 (43%) parents printed out, signed, and returned completed paper consent forms to school. Among the 188 adolescents attending the five schools that sent both e-mail and paper notifications, 152 (81%) consented online (p < 0.001 compared with e-mail-only notifications) and 36 (19%) returned completed paper consent forms.

## Discussion

### Summary of key findings

To the best of our knowledge, this is the first review to assess the available evidence on the use of electronic consent (e-consent) as a means of obtaining informed consent for school-age vaccinations. While there has been growing interest on the use of e-consent in healthcare and other research areas, there appears to be a paucity of evidence-based studies focused specifically on e-consent for school-aged vaccinations. The review findings appear to suggest a preference for e-consent systems over traditional paper-based consent by parents of school children. Parents interviewed found the system easy to use, which resulted in better engagement with school-based vaccination programmes. However, some parents and school staff found the transition from traditional paper consent to e-consent difficult to adapt to. Language barriers, practical issues with the new system, and concerns over lack of trust in electronic data, were among the reasons given for lack of engagement with the system.

The review findings showed that e-consent did not improve the outcome of consent or vaccination uptake. Again, challenges encountered in transitioning from paper to electronic consent, was highlighted as a possible cause. According to Chantler et al., (2020), issues with information technology functionality, and limited staff training and engagement of schools, hampered the transition process. The authors suggested that new technologies, like e-consent, would need to be used for more than a year before they can show any benefits and become incorporated into routine practice. Footer and Foster (2022) attributed external factors like the COVID-19 pandemic for lower vaccination uptake. The study authors theorised that a combination of government guidelines on isolation and lack of public trust in vaccines due to widespread misinformation about the COVID vaccine, was responsible for lower vaccination uptake.

Mixed findings were reported on the impact of e-consent on consent form return. One study reported lower consent form returns in schools issued with e-consent forms compared to schools issued with paper forms, while the other study reported a significantly higher return rate for children issued with e-consent forms. As this outcome was reported by only two studies with varying results, firm conclusions cannot be made on the impact of e-consent on consent form return.

Our review findings are in keeping with other evidence reviews on e-consent (Chen et al., 2020, Chimonas et al., 2023, Obaidi et al., 2024). These studies found that the number of research publications on e-consent were relatively low but emergent, which confirms our finding of the paucity of high-quality published studies on this topic. When used in clinical care or healthcare settings, improvements were reported in efficiency and user preference (Chimonas et al., 2023, Obaidi et al., 2024). However, replacing paper consent with e-consent for research was met with concern over privacy breeches, data misuse, and anonymity (Chen et al., 2020). Further research on the consequences of transitioning from paper to e-consent are therefore recommended.

### Strengths and limitations of this rapid review

Due to time and resource constraints, an abbreviated systematic review methodology was utilised in this review. This included the use of a systematic approach to identify, review, and synthesise study findings in compliance with PRISMA guidelines. An extensive literature search using a wide range of search terms and databases was also conducted. Only studies with study designs incorporating a comparative element, as well as those whose findings are generalisable to the UK, were included. The quality of included studies was assessed using critical appraisal tools well suited for the study designs of included studies.

This review is limited by the inclusion of only studies published in English language and the absence of grey literature searches. Whilst it is possible that additional eligible publications may have been missed, a thorough and transparent attempt was made to capture all relevant publications with minimal risk of bias.

### Strengths and limitations of the available evidence

All included studies had clear aims, objectives and outcome measures. Studies were mostly quantitative, but also contained qualitative aspects for better understanding of the subject. Included studies were conducted in the UK and USA, therefore, aiding the generalisability of findings to the UK context. Except for one study that was focused on elementary school children, the majority of studies focused on adolescent children in secondary school.

Only four studies were eligible for inclusion in this review, highlighting a paucity of research evidence on e-consent. Some outcomes were reported by so few studies - in some cases, one study, making it difficult to draw definitive conclusions. In some pilot studies, the e-consent intervention had not been implemented long enough for a meaningful impact assessment. Several evidence gaps were identified, notably an absence of studies assessing the impact of e-consent on vaccine equity and access. Similarly, none of the studies explored the cost-effectiveness and quantified time savings of e-consent systems or focused on DPT, MenACWY, and MMR vaccinations. Furthermore, none of the included studies explored the characteristics of parents who consented using the e-consent process or targeted children not in mainstream education.

### Implications for policy and practice

This rapid review highlights the potential benefits of e-consent over traditional paper consent systems for school-age vaccinations. Research evidence on the effectiveness of e-consent processes is currently very much emergent. However, more high-quality research is needed to inform the use of these processes for obtaining informed consent for vaccinations. Further research focusing on the consequences of transitioning from paper to e-consent, such as digital exclusion, vaccine inequity, as well as language considerations is needed to inform policy and practice.

## Conclusion

There is currently growing interest on the use of e-consent as a means of obtaining informed consent for school-aged vaccinations. However, there is limited published evidence from the small number of studies identified. When compared to paper consent, e-consent forms were found to be easier to use and quicker to complete but did not have any impact on vaccination uptake. Transitioning from paper to e-consent systems was found to be difficult for some parents and adolescents. Any change in process may take a while to embed and a decrease may be seen before its true impact is observed. Further evaluations of the benefits and consequences of e-consent are required to inform its use in vaccination processes.

## Data Availability

All data produced in the present study are available upon reasonable request to the authors

## Appendix 1 Search strategy

1 (child* or school* or teen* or adolescen* or youth* or young person* or young people or girl* or boy*).ti,ab.

2 exp Schools/

3 (school or schools).ti,ab.

4 1 or 2 or 3

5 (informed consent or informed decision or consent).ti,ab.

6 ((improv* or optimi*) adj2 consent).ti,ab.

7 (online consent or online informed consent or electronic consent or electronic informed consent or digital consent or digital informed consent or e-consent or econsent or consent by email).ti,ab.

8 ((computer or tablet or digital or electronic or electronic device or multimedia or smartphone or online systems or mobile applications or apps or internet) adj2 consent).ti,ab.

9 5 or 6 or 7 or 8

10 exp Vaccination/ or exp Immunization Programs/ or exp Immunization/

11 (immunis* or immuniz* or vaccina*).ti,ab.

12 exp Diphtheria-Tetanus-Pertussis Vaccine/

13 exp Papillomavirus Vaccines/

14 exp Meningococcal Vaccines/

15 exp Influenza Vaccines/

16 (((cervical cancer or diphtheria or DtaP or DTP or HPV or measles or MenC or MenACWY or meningitis or Meningococcal or Neisseria meningitidis or papillomavirus or pertus* or rubella or rubeola or td?ipv or tetanus or wart virus or whoop*) adj2 immunis*) or immuniz* or vaccina*).ti,ab.

17 10 or 11 or 12 or 13 or 14 or 15 or 16

18 4 and 9 and 17

## Notes

### Competing Interest Statement

The authors have declared no competing interest.

### Funding Statement

This study did not receive any funding

